# Experience with open schools and preschools in periods of high community transmission of COVID-19 in Norway during the academic year of 2020/2021

**DOI:** 10.1101/2021.11.16.21265186

**Authors:** Sara Stebbings, Torill Alise Rotevatn, Vilde Bergstad Larsen, Pål Surén, Petter Elstrøm, Margrethe Greve-Isdahl, Tone Bjordal Johansen, Elisabeth Astrup

**Author notes:** **Corresponding author:** Sara Stebbings, Division of Infection Control and Environmental Health, Norwegian Institute of Public Health, Postboks 222, Skøyen, N-0213. Equal contributors (Tone Bjordal Johansen, Elisabeth Astrup).

## Abstract

**Background:** Schools and preschools have largely remained open in Norway throughout the pandemic, with flexible mitigation measures in place. This contrasts with many other high-income countries that closed schools for long periods of time. Here we describe cases and outbreaks of COVID-19 in schools and preschools during the academic year 2020/2021, to evaluate the strategy of keeping these open with infection prevention control measures in place.

**Methods:** In this descriptive study, the Norwegian Institute of Public Health initiated systematic surveillance for COVID-19 cases and outbreaks in schools and preschools in October 2020. Data was compiled from the national outbreak alert system VESUV, municipality websites, and media scanning combined with the national emergency preparedness register Beredt C-19. An outbreak was defined as ≥ 2 cases among pupils or staff within 14 days at the same educational setting. Settings were categorized as preschool (1-5-years), primary school (6-12-years), lower secondary school (13-15-years) and upper secondary school (16-18-years). We reported the incidence rate among preschool and school-aged pupils and gave a descriptive overview of outbreaks and included cases per educational setting.

**Results:** During the whole academic year, a total of 1203 outbreaks in preschools and school settings were identified, out of a total of 8311 preschools and schools nationwide. The incidence of COVID-19 in preschool- and school-aged children and the rates of outbreaks in these settings largely followed the community trend. Most of the outbreaks occurred in primary schools (40%) and preschools (25%). Outbreaks across all settings were mostly small (median 3 cases, range 2 to 72), however, 40 outbreaks (3% of total) included 20 or more cases. The larger outbreaks were predominantly seen in primary schools (43%).

**Conclusions:** We observed few large outbreaks in open schools and preschools in Norway during the academic year of 2020/2021, also when the Alpha variant was predominant. This illustrates that it is possible to keep schools and preschools open even during periods of high community transmission of COVID-19. Adherence to targeted IPC measures adaptable to the local situation has been essential to keep educational settings open, and thus reduce the total burden on children and adolescents.

## Background

COVID-19 has had a disastrous effect on the access to school for millions of children globally. The United Nations estimated that more than 168 million school-children missed out on learning in class during the first year of the pandemic (1). In addition, around one in seven pupils globally (214 million) missed more than three-quarters of their in-person learning, and many more have had their education partially or entirely disrupted. Education is one of the strongest predictors of a population’s health and prosperity, and children have a fundamental right to attend school (2, 3).

Quite early in the pandemic, it became evident that children had lower rates of hospitalization and death caused by COVID-19 than all other age groups (4). This also holds true for variants of concern such as the Alpha and Delta variants (5, 6). Children are less susceptible and probably transmit onward to a lesser degree than other age groups, although their role in the transmission of SARS-CoV-2 is still debated (7).

Cases and outbreaks in schools and preschools seem to reflect local incidence levels (8). General mitigation measures in society influence the number of cases seen in schools. In Norway, schools and preschools closed during the first wave of the pandemic during the course of spring 2020, but re-opened from late April 2020 with infection prevention and control (IPC) guidelines in place (9).

As schools reopened, there was a need for more flexible IPC-measures to allow for a response that was better adapted to different incidence levels, whereas low incidence levels allow for normal class size and in-person education. A flexible traffic-light model (TLM) adaptable to the local epidemiological situation and age group, was therefore developed (figure 1). Norwegian schools and preschools implemented the model and have largely remained open throughout the pandemic, with only short-term closures (full-time digital teaching), mainly in secondary schools, as a response to local outbreaks. This also applied during the second and third waves in contrast to many other countries where schools and preschools were closed for longer periods (10).

**Figure 1.**
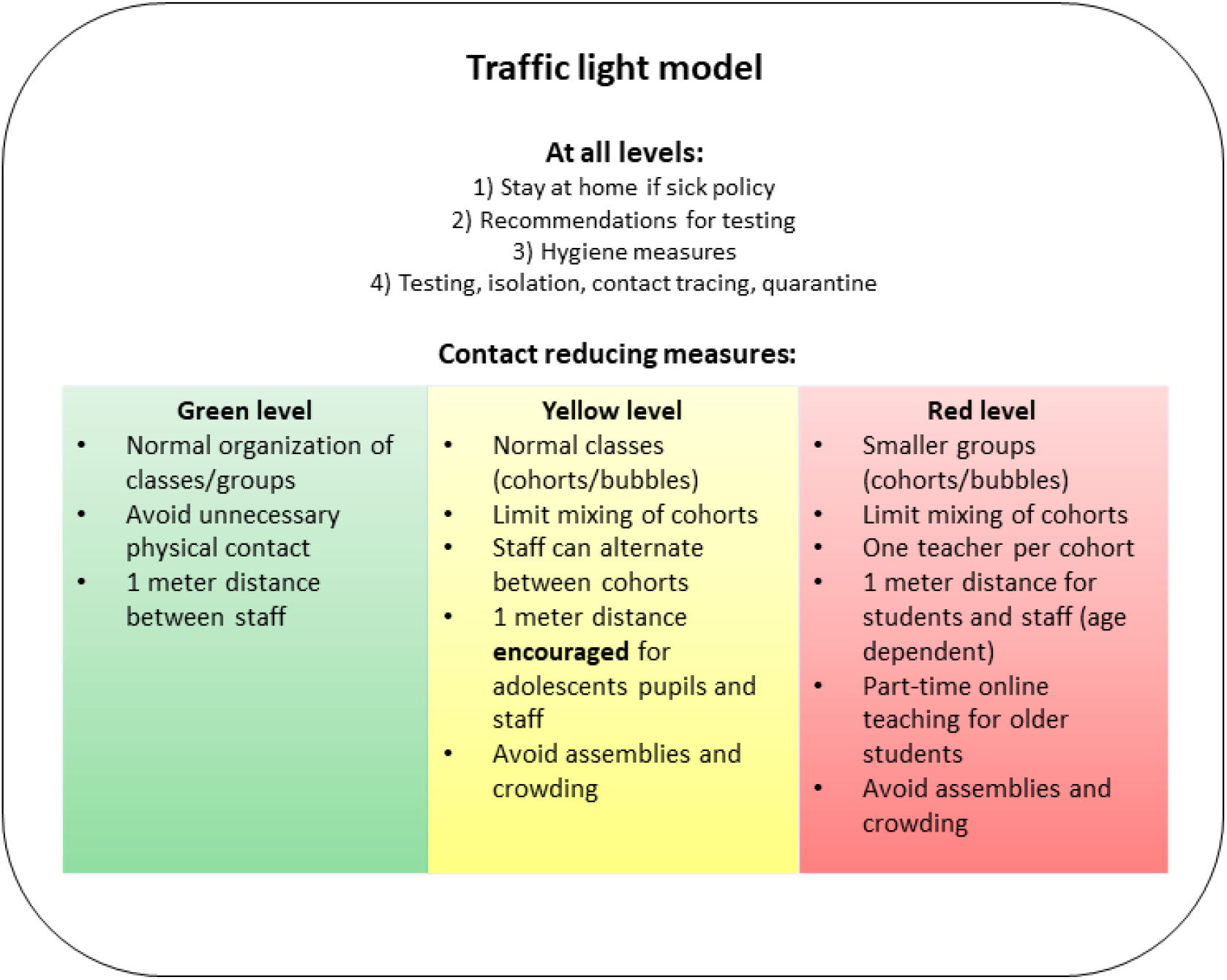
Traffic light model, main features. Notes: The full IPC guidelines are published by the national educational authorities (11).

The TLM has three levels of measures (figure 1): green (normal situation), yellow (intermediate, cohorts with normal class sizes), and red (distance requirements, smaller cohorts/ groups). The main difference between the levels is the contact reducing measures. Red level implies stricter distance requirements and smaller groups, which means a combination of in-person and digital education for older pupils (hybrid model). At yellow level class sizes are normal, but classes are considered cohorts and should not mix. Face masks have been in very limited use for staff and older students only and restricted to areas with high ongoing community transmission.

The TLM was implemented at yellow (intermediate) level nationwide from June 2^nd^, 2020. Red level has been implemented in municipalities with high community transmission when needed. Green level was not applied during the study period.

The aim of this study is to describe cases and clusters/outbreaks of COVID-19 in Norwegian preschools and schools, to evaluate the strategy of keeping these settings open with IPC measures in place. The period covered most of the academic year 2020-2021, including both the second wave, dominated by the original virus variant, and the third wave, dominated by the Alpha variant.

## Methods

### Data sources

In this descriptive study, the Norwegian Contact Tracing Team (NCTT) initiated systematic surveillance of COVID-19 cases and clusters (pupils and staff) in schools and preschools in October 2020. We collected data from the national outbreak alert system VESUV where all outbreaks, suspected or confirmed, are reported to The Norwegian Institute of Public Health (NIPH) (12). Reporting is mandated by law and conducted by various official health authorities. In addition, regular screening of the twenty largest municipality websites, as well as media scanning was performed.

In events where no information was available online, or more detailed information was needed, we contacted municipal health authorities and local contact tracing teams. We obtained the following variables on clusters in schools and preschools: date of onset, educational setting and the number of cases included in outbreaks distributed by pupils and staff.

Further, data was obtained from the national emergency preparedness register (Beredt C-19) to present the weekly number of confirmed COVID-19 cases in Norway during the study period (13). From within Beredt C-19, data was compiled from the Norwegian Surveillance System for Communicable Diseases (MSIS) and the National Population Register (age and demographic information) (14). COVID-19 is a notifiable disease in Norway and all cases are registered in MSIS.

### Study population and educational settings

The study period was set from 28^th^ September 2020 (week 40) to 6^th^ June 2021 (week 22). In this study we define the second and third wave of COVID-19 in Norway as October 2020 to January 2021 (week 43-week 2) and mid-February to mid-April (week 7-week 16), respectively. A case was defined as a child/pupil or staff who tested positive for SARS-CoV-2 (PCR or antigen rapid test) and had physically attended preschool or school during their infectious period. Infectious period was defined as 48 h before symptom onset (48 h before test date if asymptomatic) until the end of the isolation period. All staff members were included, regardless of role (educational or non-educational). Details on educational settings in Norway are given in table 1 (15, 16).

**Table 1.** Characteristics of the educational system in Norway.

|  | Preschool | Primary school | Lower secondary school | Upper secondary school |
| --- | --- | --- | --- | --- |
| Age group | 1-5 years | 6-12 years | 13-15 years | 16-18 years |
| Number of institutions | 5,620 | 2276* |  | 415 |
| Number of children/pupils | 272.264 | 635.497* |  | 188.214 |
| Number of staff | 95.546 | 115.205* |  | 39.681 |
| Children/pupils per educational staff | 5.7 | 16* |  | NA |

\*Total number for primary and lower secondary school (our data does not separate between these school levels).

### Cases and outbreaks

We reported outbreaks per educational setting. An outbreak was defined as two or more positive cases among pupils and/or staff within 14 days at the same educational setting. An outbreak was considered over when no new cases were reported at the facility within 14 days.

Information regarding the epidemiological link of cases was not always available. Hence, we considered clusters in the same school/preschool to be *possible outbreaks*. For the sake of simplicity, the term outbreak was used for both possible and confirmed outbreaks.

The date of onset of the outbreak was defined as the date of symptom onset for the index case if available. If not available, we used the date an outbreak was reported.

### Statistical analysis

The 7-day incidence rate of newly reported COVID-19 cases per 100,000 inhabitants was plotted by week (week 40, 2020, and week 22, 2021) by age group for preschool and school-aged pupils, as well as the total number of confirmed cases in the population. Further, we visualized the weekly number of outbreaks by educational setting to observe the trends over time. School holidays during the time of study were also included. Data management was performed using R Statistical Software (version 3.6.2).

### Ethics

NIPH has under legal permission through the Communicable Disease Act and MSIS the right to process patient confidential information for national surveillance of communicable diseases and outbreak investigation (14). The establishment of an emergency preparedness register (Beredt C-19), forms part of the legally mandated responsibilities of NIPH during epidemics (13). Beredt C19 was established under Section 2–4 of the Health Preparedness Act. NIPH has conducted a data protection impact assessment of the register. In addition, a study protocol which covers the current work, was granted approval by the Ethics Committee of South-East Norway (28^th^ June 2021, #240725).

## Results

### Number of cases

The incidence of reported COVID-19 cases for preschool- and school-aged children largely followed the trend in the community (figure 2). During the 2020/2021 academic year, the second wave in Norway started in October after the autumn school holiday (week 40 or 41) and peaked in November (week 45-46). Further, an increase in infection rates was seen following the Christmas holiday (week 53-1, 2020/2021). In this period, a larger proportion of adolescents in lower and upper secondary schools tested positive for COVID-19 compared to younger children. The third wave started in late February around the winter school holiday (week 8 or 9) and peaked in March (week 11). A larger proportion of the preschool- and primary school-aged children were infected than in previous waves, even though cases among young adults and adolescents dominated. This coincided with the introduction and subsequent dominance of the Alpha variant in Norway. Distinct peaks were seen in infection rates for the oldest adolescents (upper secondary school) in May and early June (week 18-22).

**Figure 2.**
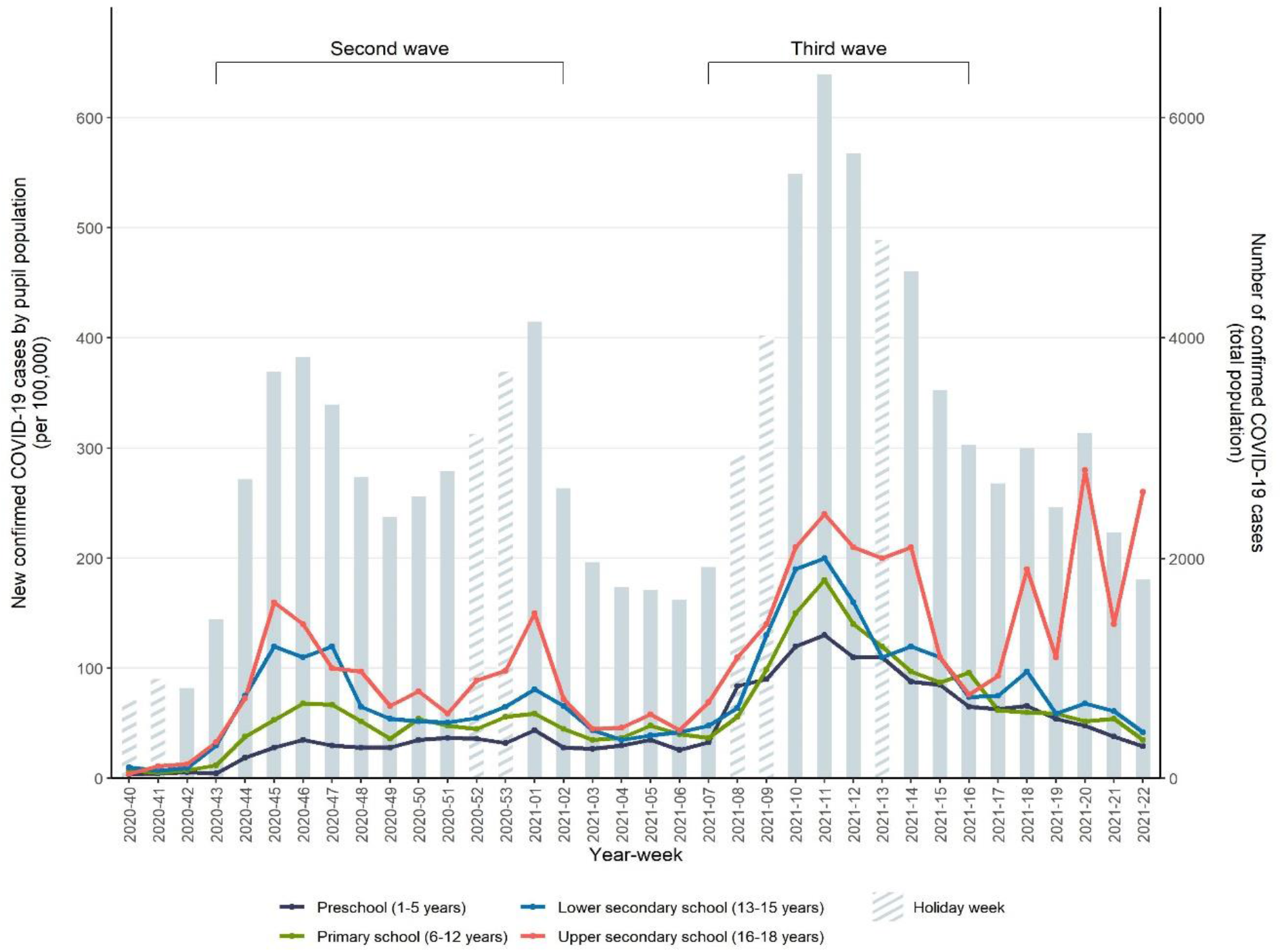
Seven-day incidence rate of reported COVID-19 cases per 100,000 inhabitants by week and education level and total number.

### Outbreaks

The NCTT surveillance registered 2866 COVID-19 related events in preschools and schools during the study period. Of these, 1189 (41%) entries were excluded as they only included a single case, leaving 1677 outbreaks. Of these, another 474 (28%) outbreaks were excluded as they lacked information on the number of cases. These outbreaks were mainly reported through municipality websites (85%) during and after the third wave (week 7 – week 22). The remaining 1203 outbreaks were included in the analysis (Table 2).

**Table 2.** Summary of outbreaks and cases in preschools and schools reported or notified to the NCTT.

|  | Preschools | Primary | Combined<sup>†</sup> | Lower secondary | Upper secondary | Total |
| --- | --- | --- | --- | --- | --- | --- |
| **Outbreaks** (n (%)) | 298 (25) | 457 (40) | 33 (3) | 190 (16) | 225 (19) | 1203 |
| Pupils only | 58 (10) | 230 (40) | 18 (3) | 125 (22) | 143 (25) | 574 |
| Staff only | 51 (71) | 13 (18) | 0 (0) | 1 (1) | 7 (10) | 72 |
| ≥ 20 cases | 8 (20) | 17 (43) | 2 (5) | 5 (13) | 8 (20) | 40 |
| **Cases** (n (%)) | 1447 (22) | 2446 (37) | 255 (4) | 1018 (16) | 1376 (21) | 6530 |
| Pupils/children | 799 (16) | 1917 (38) | 214 (4) | 894 (18) | 1208 (24) | 5032 |
| Staff members | 648 (43) | 528 (35) | 41 (3) | 124 (8) | 157 (10) | 1498 |
| Infected staff per infected pupils/children | 0.8 | 0.3 | 0.2 | 0.1 | 0.1 | 3.4 |
| Mean (SD*), per outbreak | 4.9 (4.5) | 5.4 (5.6) | 7.7 (7.6) | 5.4 (5.8) | 6.1 (8.2) | 5.4 (6.0) |
| Median (IQR**), per outbreak | 3 (2-5) | 3 (2-6) | 4 (3-10.5) | 3 (2-6) | 4 (2-7) | 3 (2-6) |
| Range | 2-30 | 2-52 | 2-34 | 2-55 | 2-72 | 2-72 |

<sup>†</sup> Combined primary and lower secondary schools
\*Standard deviation
\*\*Interquartile range

Most outbreaks were reported in primary schools (n= 457), followed by preschools (n= 298), upper secondary (n= 225), lower secondary (n= 190) and combined schools (n= 33), out of a total of 8311 preschools and schools nationwide.

Outbreaks fluctuated across all settings throughout the study period with the highest peaks during the second and third wave in week 45, 2020 (n= 58) and week 11, 2021 (n= 69) (figure 3). This largely followed the community trend. In general, a declining trend in the number of outbreaks was observed during holiday weeks, followed by an increasing trend shortly after.

**Figure 3.**
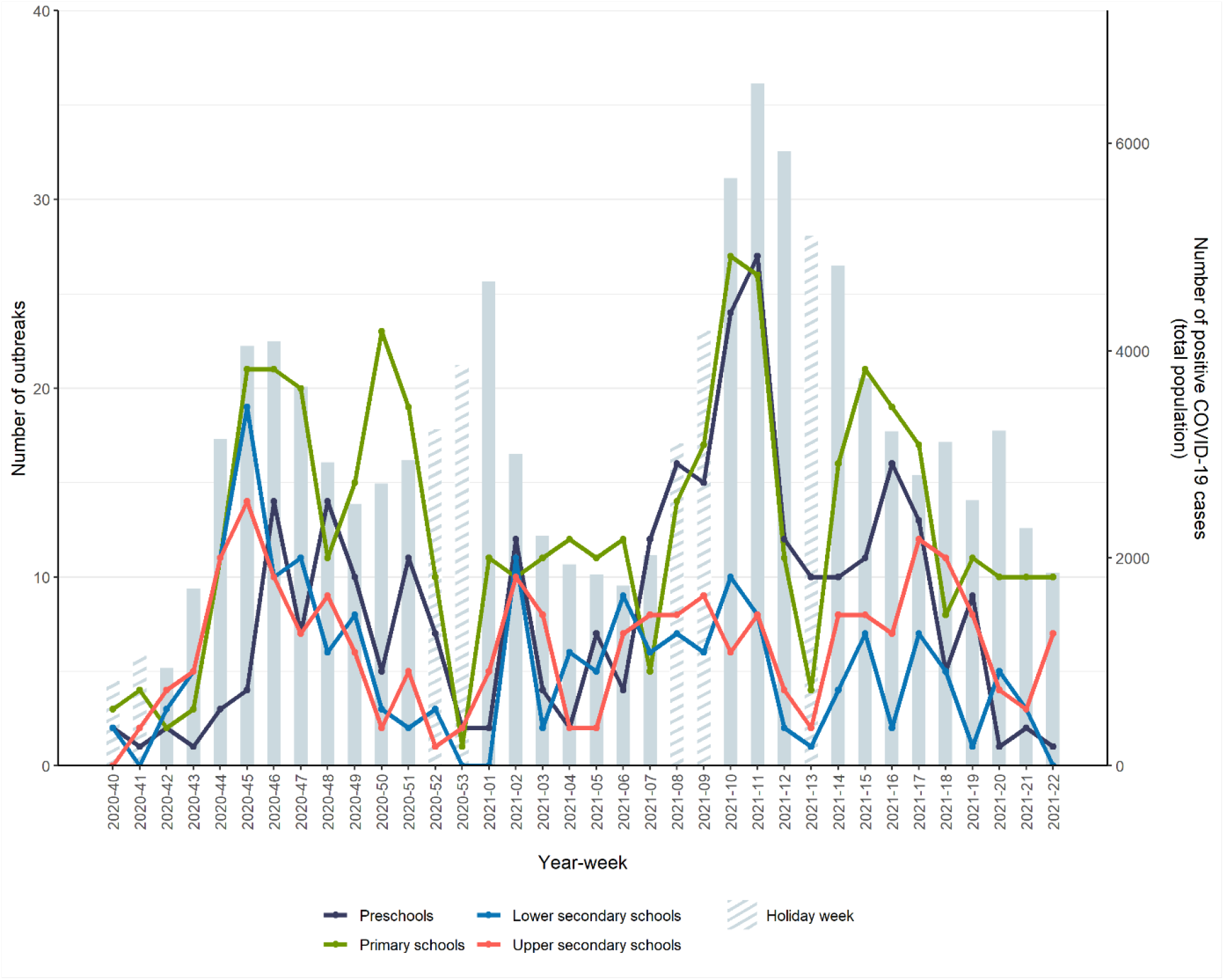
Number of outbreaks by setting and week from week 40, 2020 to week 22, 2021.

In total, there were 6530 cases included in the 1203 outbreaks, of whom 5032 were pupils and 1498 were staff members (table 2). In almost half of the outbreaks, cases were limited to pupils only (n= 574, 48%), most frequently observed in primary and upper secondary schools (n= 230 and n= 143, respectively). This contrasted with outbreaks involving staff only (n=72), where the majority was observed in preschools (n=51, 71 %). This was also reflected by the higher ratio of cases among staff versus children/pupils observed in preschools (0.8) followed by primary schools (0.3).

Most cases were reported from primary schools and preschools, with 2446 (37%) and 1447 (22%) cases. The highest number of cases among pupils was observed in primary schools (n= 1917, 38%) followed by upper secondary schools (n=1208, 24%). Cases among staff members were more common in preschools (n= 648, 43%) and primary schools (n= 528, 35%).

We observed mostly small outbreaks, however with large variation (median= 3, range= 2-72). There were in total 40 outbreaks (3%) with ≥ 20 cases. The majority of these occurred in primary schools (n=17, 43%). Most larger outbreaks across both schools and preschools occurred during the third wave dominated by the Alpha variant (n=18, 45%).

## Discussion

In Norway, unlike most other high-income countries, schools at all levels remained open throughout the academic year 2020/2021. In areas and periods with high transmission, strict IPC measures (such as red level in the TLM) were implemented to limit transmission and outbreaks. A substantial number of cases and outbreaks were observed in schools and preschools during this period, however, only a small fraction of the outbreaks included more than twenty cases.

In the second wave starting autumn 2020, the incidence followed the same trend in all age groups, with most cases seen in older pupils. In the third wave, which coincided with the introduction of the Alpha variant, incidence in children in the youngest age groups for the first time came close to the incidence rate among older pupils. A similar trend was observed internationally (6). Although the incidence was highest among adolescents, the highest number of outbreaks were observed in primary schools and preschools. Several factors may have contributed to this. First, there are more preschools and primary schools than secondary schools in Norway, as well as more children/pupils attending these facilities (12 versus 6 age cohorts). Second, stricter IPC measures (red level) were implemented in secondary, and especially in upper secondary schools, during periods with high transmission rates.

As the third wave subsided, there was a sudden increase in cases in upper secondary schools, including some of the largest outbreaks during the study period. This coincided with graduation celebrations, characterized by intense socializing over several weeks. Contact tracing revealed that the majority of these cases were likely infected outside schools. During this period, affected municipalities reported increasing pandemic fatigue in this group and difficulties with complying to IPC measures, a phenomenon also observed elsewhere (17).

The higher ratio of cases among staff versus children/pupils in preschools and primary schools may in part be explained by the higher number of staff per child working in these settings. Additionally, there is a naturally closer contact both in-between staff and between staff and children as these settings involve care for young children. In fact, contact between adults and children in preschool resembles the contact in households. In a recent Norwegian study, transmission within households was higher from younger children than from older children and adolescents, again explained by the same contact pattern between small children and parents (18). The nature of working with care for young children illustrates the importance of IPC measures tailored to the specific setting. In addition, it is essential to provide vaccination to staff.

Closing schools and loss of in-person teaching have well-documented profound negative consequences on learning progress and mental health (19, 20, 21). The negative consequences are greater in more deprived families, thus worsening health and educational inequalities. Existing evidence on the effects of school closures in reducing community transmission of SARS-CoV-2 is uncertain and varies from no to some effect. Several studies have shown that school reopening with IPC measures in place did not increase community transmission (22, 23). Data from Norway indicate that closure gives no added effect compared to open schools with strict IPC measures (red level) in place in areas with high incidence levels (24, 25). In addition, a recent study from the United States suggests that opening schools for in-person learning with appropriate IPC measures in place leads to minimal increase in community incidence of SARS-CoV-2 (26).

The incidence in children and adolescents largely followed the trend in the community. This supports that schools can remain open with mitigation measures, without profoundly affecting community transmission. School closure may in fact not lead to an actual reduction in contact among pupils, depending on the level of social mixing outside of school. In our experience, we have observed an increase in cases and outbreaks after school holidays. Reports from contact tracing teams indicate that much of this transmission was linked to social activities. In addition, some of the largest outbreaks were observed among adolescents and associated with social settings outside school. These outbreaks would not have been avoided by closing schools. Long-term closure may also lead to increased spontaneous social contact in other arenas, where no IPC measures are in place. An increasing number of studies indicate that closing schools have limited effect on the transmission of COVID-19. As long as closing schools is not evidence-based and the negative effects are so profound, we should always strive to limit school closures to an absolute minimum (27, 28, 29).

In order to keep schools open, it is important to implement IPC measures. However, IPC measures are intrusive and limit both education and social activities in school. Compliance may be difficult for children. Thus, measures should always be adjusted according to the local situation and the use of strict measures should be limited. Non-pharmaceutical interventions for the society at large are equally essential to reduce community transmission (6). This has been systematically implemented in Norway. Close cooperation between public health and educational authorities and stakeholders is important to implement the guidelines and ensure good quality education for children during health threats like the COVID-pandemic. How to prioritize restrictions between groups and interests is in large part a political decision. The Norwegian government early declared that children and adolescents should be prioritized, thus there has been a political will to keep schools and preschools open, at the expense of other sectors. With increasing vaccine coverage in the population, the need for strict IPC measures in schools and preschools will subside.

The strength of this study lies in the high-quality registry data from Beredt C-19, allowing detailed information regarding incidence in the different age groups. This, in combination with multiple other data sources, provided completeness of the data and enabled us to present more detailed results regarding outbreaks in preschools and schools.

The manual data collection done in the NCTT surveillance has some limitations and is resource-intensive. First, reporting outbreaks in the national outbreak system VESUV is mandatory. However, we are aware that not all outbreaks were notified, particularly during the transmission peaks throughout the study period. Secondly, the quality of information available through municipality websites and news media varied. During periods with high transmission, the reports may have been less accurate due to capacity challenges. Thirdly, some outbreaks were excluded due to missing information regarding the number of cases, whereas the majority occurred during and after the third wave. These factors may have led to an underreporting of outbreaks during the study period. However, as NIPH is regularly consulted by municipalities when they experience larger outbreaks, we believe the outbreaks we may have missed were mainly small, with limited need for comprehensive measures.

In all, we believe that combining different data sources provided sufficient and adequate data to allow us to expand existing information regarding transmission and outbreak trends in schools and preschools during the academic year of 2020/2021.

## Conclusion

Schools and preschools in Norway remained open during the academic year of 2020/2021. We observed few large outbreaks in educational settings during this period, also during the third wave dominated by the Alpha variant. This illustrates that it is possible to keep schools and preschools open even during periods of high community transmission of COVID-19. Large outbreaks are not necessarily a result of in-school transmission, and in some of the largest outbreaks a substantial number of cases were probably associated with transmission in social settings outside school. These would not have been avoided by closing schools. Adherence to targeted IPC measures adaptable to the local situation has been essential to keep educational settings open, and thus reduce the total burden on children and adolescents. It is still important to monitor the spread in schools, evaluate implemented measures, and adjust to the local situation with emerging virus variants and to the increasing vaccine coverage.

## Data Availability

All data produced in the present study are available upon reasonable request to the authors.

## Abbreviations

IPC: Infection prevention and control
MSIS: Norwegian surveillance system for communicable diseases
NCTT: Norwegian contact tracing team
NIPH: Norwegian Institute of Public Health
TLM: Traffic Light Model

## Declarations

### Ethics approval and consent to participate

Our research was granted approval by the Ethics Committee of South-East Norway.

### Consent for publication

Not applicable.

### Availability of data and materials

The datasets used and/or analyzed during the current study are available from the corresponding author on reasonable request.

### Authors’ contribution

All authors designed and conceptualized the study. SS coordinated the study and conducted the data collection for the NCTT surveillance. TR and VB performed data analysis and helped to interpret the data. SS, TB and EA equally contributed to write the first draft of the manuscript, with critical reviews from TR, VB, PE, MG and PS. All authors have read and approved the final manuscript.

## Acknowledgements

We would like to thank our colleague at NIPH Idunn Aune Forland for contributing to the data collection for the NCTT surveillance.

## References

1. The United Nations International Children’s Emergency Fund. COVID-19 and School Closures: One year of education disruption [Internet]. UNICEF;2021 [cited 2021 June 11]. Available from: https://data.unicef.org/resources/one-year-of-covid-19-and-school-closures.

2. United Nations Commission on Human Rights. Convention on the Rights of the Child. Geneva: UN Commission on Human Rights; 1990 March.

3. Viner RM, Russell SJ, Croker H, Packer J, Ward J, Stansfield C, et al. School closure and management practices during coronavirus outbreaks including COVID-19: a rapid systematic review. Lancet Child Adolesc Health. 2020;4(5):397–404.

4. Viner RM, Mytton OT, Bonell C, Melendez-Torres GJ, Ward J, Hudson L, et al. Susceptibility to SARS-CoV-2 Infection Among Children and Adolescents Compared With Adults: A Systematic Review and Meta-analysis. JAMA Pediatrics. 2021;175(2):143–56.

5. Gaythorpe KAM, Bhatia S, Mangal T, Unwin HJT, Imai N, Cuomo-Dannenburg G, et al. Children’s role in the COVID-19 pandemic: a systematic review of early surveillance data on susceptibility, severity, and transmissibility. Sci Rep. 2021;11(1):13903.

6. European Centre for Disease Prevention and Control. COVID-19 in children and the role of school settings in transmission - second update [Internet]. Stockholm: ECDC; 2021 [updated 8 July 2021; cited 2021 Oct 19]. Available from: https://www.ecdc.europa.eu/sites/default/files/documents/COVID-19-in-children-and-the-role-of-school-settings-in-transmission-second-update.pdf.

7. Public Health England. Investigation of novel SARS-CoV-2 variant - Variant of Concern 202012/01 [Internet]. London: Public Health England; 2021 [cited 2021 June 24]. Available from: https://assets.publishing.service.gov.uk/government/uploads/system/uploads/attachment_data/file/959426/Variant_of_Concern_VOC_202012_01_Technical_Briefing_5.pdf.

8. European Centre for Disease Prevention and Control. COVID-19 in children and the role of school settings in transmission - first update. [Internet]. Stockholm: ECDC; 2020 [cited 2021 June 10]. Available from: https://www.ecdc.europa.eu/sites/default/files/documents/COVID-19-in-children-and-the-role-of-school-settings-in-transmission-first-update_1.pdf.

9. Johansen TB, Astrup E, Jore S, Nilssen H, Dahlberg BB, Klingenberg C, et al. Infection prevention guidelines and considerations for paediatric risk groups when reopening primary schools during COVID-19 pandemic, Norway, April 2020. Eurosurveillance. 2020;25(22):2000921.

10. United Nations Educational, Scientific and Cultural Organization. Education: From disruption to recovery [Internet]. Paris: UNESCO; 2021 [cited 2021 June 13]. Available from: https://en.unesco.org/covid19/educationresponse.

11. Utdanningsdirektoratet. Beredskap: Veiledere om smittevern i barnehager og skoler [Internet]. Oslo: Utdannignsdirektoratet; 2021 [cited 2021 Oct 1]. Available from: https://www.udir.no/kvalitet-og-kompetanse/sikkerhet-og-beredskap/informasjon-om-koronaviruset/smittevernveileder/.

12. Guzman-Herrador B, Vold L, Berg T, Berglund TM, Heier B, Kapperud G, et al. The national web-based outbreak rapid alert system in Norway: eight years of experience, 2006–2013. Epidemiology and Infection. 2016;144(1):215–24.

13. Norwegian Institute of Public Health (NIPH). Emergency preparedness register for COVID-19 (Beredt C19) Oslo: NIPH; 2020 [updated 2021 Aug 8; cited 2021 May 5]. Available from: https://www.fhi.no/en/id/infectious-diseases/coronavirus/emergency-preparedness-register-for-covid-19/.

14. Forskrift om Meldingssystem for smittsomme sykdommer (MSIS-forskriften) 2003 [LOV-2021-10-08-2958. Available from: https://lovdata.no/dokument/SF/forskrift/2003-06-20-740.

15. Statistics Norway. Ansatte i barnehage og skole [Internet]. Oslo: Statistics Norway; 2021 [updated 2021 June 22; cited 2021 June 15]. Available from: https://www.ssb.no/utdanning/barnehager/statistikk/ansatte-i-barnehage-og-skole.

16. Utdanningsdirektoratet. Statistikk [Internet]. Oslo: Utdanningsdirektoratet; 2021 [cited 2021 June 15]. Available from: https://www.udir.no/tall-og-forskning/statistikk/.

17. World Health Organization. Pandemic fatigue: reinvigorating the public to prevent COVID-19: policy framework for supporting pandemic prevention and management. Copenhagen: WHO Regional Office for Europe; 2020 [updated 2020 Nov, cited 2021 Oct 1]. Available from: https://apps.who.int/iris/bitstream/handle/10665/337574/WHO-EURO-2020-1573-41324-56242-eng.pdf?sequence=1&isAllowed=y

18. Telle K, Jørgensen SB, Hart R, Greve-Isdahl M, Kacelnik O. Secondary attack rates of COVID-19 in Norwegian families: a nation-wide register-based study. European Journal of Epidemiology. 2021;36(7):741–8.

19. Engzell P, Frey A, Verhagen MD. Learning loss due to school closures during the COVID-19 pandemic. Proc Natl Acad Sci USA.2021;118(17).

20. Larsen L, Helland MS, Holt T. The impact of school closure and social isolation on children in vulnerable families during COVID-19: a focus on children’s reactions. Eur Child Adolesc Psychiatry. 2021:1–11.

21. Viner RM, Bonell C, Drake L, Jourdan D, Davies N, Baltag V, et al. Reopening schools during the COVID-19 pandemic: governments must balance the uncertainty and risks of reopening schools against the clear harms associated with prolonged closure. Archives of disease in childhood. 2021;106(2):111–3.

22. Walsh S, Chowdhury A, Braithwaite V, Russell S, Birch J, Ward J, et al. Do school closures and school reopenings affect community transmission of COVID-19? A systematic review of observational studies. BMJ Ope. 2021;11(8).

23. National Collaborating Centre for Methods and Tools. Living Rapid Review Update 17: What is the specific role of daycares and schools in COVID-19 transmission? Canada: McMaster University; 2021 [cited 2021 Oct 1]. Available from: https://www.nccmt.ca/uploads/media/media/0001/02/e7b567a4ea25117ff577c83c9e2d8963100af547.pdf

24. Astrup E EP, Greve-Isdahl M, Johansen TB, Rotevatn TA, Surén P. Evaluering av effekt av smitteverntiltak i skoler februar-april 2020. [Internet]. Oslo: Folkehelseinstituttet; 2021 [cited 2021 May 24] Avaliable from: https://www.fhi.no/globalassets/dokumenterfiler/rapporter/2021/evaluering-av-effekt-av-smitteverntiltak-i-skoler-februar-april-2021-rapport-2021.pdf

25. Rotevatn TA, Larsen VB, Bjordal Johansen TK, Astrup E, Surén P, Greve-Isdahl M, et al. Transmission of SARS-CoV-2 in Norwegian schools: A population-wide register-based cohort study on characteristics of the index case and secondary attack rates. medRxiv. 2021:2021.10.04.21264496.

26. Ertem Z, Schechter-Perkins EM, Oster E, van den Berg P, Epshtein I, Chaiyakunapruk N, et al. The impact of school opening model on SARS-CoV-2 community incidence and mortality. Nature Medicine. 2021.

27. Fukumoto K, McClean CT, Nakagawa K. No causal effect of school closures in Japan on the spread of COVID-19 in spring 2020. Nature Medicine. 2021.

28. Juutinen A, Sarvikivi E, Laukkanen-Nevala P, Helve O. Closing lower secondary schools had no impact on COVID-19 incidence in 13-15-year-olds in Finland. Epidemiol Infect. 2021;149:e233.

29. Lewis SJ, Munro APS, Smith GD, Pollock AM. Closing schools is not evidence based and harms children. BMJ. 2021;372:521.

